# Mechanisms underlying exercise intolerance in Long COVID: an accumulation of multi-system dysfunction

**DOI:** 10.1101/2023.12.07.23299429

**Authors:** Alexandra Jamieson, Lamia Al Saikhan, Lamis Alghamdi, Lee Hamill Howes, Helen Purcell, Toby Hillman, Melissa Heightman, Thomas Treibel, Michele Orini, Robert Bell, Marie Scully, Mark Hamer, Nishi Chaturvedi, Hugh Montgomery, Alun D. Hughes, Ronan Astin, Siana Jones

## Abstract

The pathogenesis of exercise intolerance and persistent fatigue which can follow an infection with the SARS-CoV-2 virus (’Long COVID’) is not fully understood.

Cases were recruited from a Long COVID clinic (N=32; 44±12y; 10(31%)men), and age/sex-matched healthy controls (HC) (N=19; 40±13y; 6(32%)men) from University College London staff and students. We assessed exercise performance, lung and cardiac function, vascular health, skeletal muscle oxidative capacity and autonomic nervous system (ANS) function. Key outcome measures for each physiological system were compared between groups using potential outcome means(95% confidence intervals) adjusted for potential confounders. Long COVID participant outcomes were compared to normative values.

When compared to HC, cases exhibited reduced Oxygen Uptake Efficiency Slope (1847(1679,2016) vs (2176(1978,2373) ml/min, p=0.002) and Anaerobic Threshold (13.2(12.2,14.3) vs 15.6(14.4,17.2) ml/Kg/min, p<0.001), and lower oxidative capacity on near infrared spectroscopy (τ: 38.7(31.9,45.6) vs 24.6(19.1,30.1) seconds, p=0.001). In cases, ANS measures fell below normal limits in 39%.

Long COVID is associated with reduced measures of exercise performance and skeletal muscle oxidative capacity in the absence of evidence of microvascular dysfunction, suggesting mitochondrial pathology. There was evidence of attendant ANS dysregulation in a significant proportion. These multi-system factors might contribute to impaired exercise tolerance in Long COVID sufferers.

**Key Points:**

- The pathogenesis of exercise intolerance and persistent fatigue which can follow an infection with the SARS-CoV-2 virus (’Long COVID’) is not fully understood.
- We show that Long COVID is associated with reduced measures of exercise performance in line with previous work.
- In Long COVID cases, we observed reduced skeletal muscle oxidative capacity in the absence of evidence of microvascular dysfunction, suggesting mitochondrial pathology.
- We also observed evidence of attendant autonomic nervous system (ANS) dysregulation in a significant proportion of Long COVID cases.
- These multi-system factors might contribute to impaired exercise tolerance in Long COVID sufferers.

## Introduction

Long COVID or post-acute sequelae of SARS-CoV-2 infection (PASC), is defined as the presence of persistent, often severely debilitating, symptoms beyond 12-weeks from an acute episode of COVID-19. 3.1% of the UK population self-report Long COVID symptoms [1] and it can occur irrespective of the severity of acute COVID-19 symptoms. Thus, understanding the pathogenesis of Long COVID symptoms is an important clinical challenge.

Extreme fatigue and exercise intolerance are common symptoms of Long COVID [2, 3], many studies have reported severe reductions in exercise capacity or cardiopulmonary fitness, [4, 5] however, the pathogenesis of this impairment is not fully understood. Impairments of peripheral oxygen extraction have been implicated [4, 5]. One recent study performed comprehensive assessments of skeletal muscle physiology and showed a reduction in oxidative capacity and mitochondrial enzymes [6]. However, deficits across multiple physiological systems may also contribute to exercise intolerance. Dysautonomia [7–9], anaemia [10], and a range of cardiovascular abnormalities, including right ventricular dysfunction and arrhythmias, have been described in the presence of Long COVID [11]; all of which are known to impact exercise performance.

We aim to describe, within one study, the function of several key physiological systems important for exercise performance (cardiac, pulmonary, skeletal muscle and ANS function) in people with symptoms of Long COVID compared to healthy control participants and, for clinical context, compare them to accepted normal clinical thresholds (where available from guideline documents). Using statistical modelling we also explore whether deficits in any of these systems can explain the reduction in exercise performance in people with Long COVID. We hypothesise that peripheral impairments to skeletal muscle energetics will explain deficits in aerobic capacity in Long COVID.

## Methods

The study was performed in accordance with the principles of the declaration of Helsinki and approved by the Leicester Central Research Ethics Committee. Ethical approval for recruitment of healthy individuals was granted by the University College London Research Ethics Committee. The study was registered on ClinicalTrials.gov (NCT04914754).

### Study Participants

Long COVID cases were identified by clinicians in the adult post COVID-19 clinic at University College London Hospitals NHS Foundation Trust (UCH). Patients were considered eligible for the study if they had self-reported exercise intolerance and fatigue that developed during or after an acute COVID-19 infection (confirmed by SARS-CoV-2 PCR testing), which persisted for ≥12 weeks, and which was not explained by an alternative diagnosis. Healthy adult controls were recruited from University College London (UCL) staff and students and were sex and age (5-year banding) matched to cases. Full exclusion criteria are defined in the supplementary information file (1.1). Individuals with Long COVID performed an extended version of the study protocol to assess additional measures of cardiac, muscle, vascular and autonomic function. All research procedures took place at the Bloomsbury Centre for Clinical Phenotyping (BCCP), UCL. All participants gave written informed consent.

### Participant characteristics and anthropometrics

Participant age, sex, ethnicity, co-morbidities and Long COVID symptomatology were collected by questionnaire and recorded in REDCap. Height was measured using a stadiometer (Seca217, Seca, Germany) to the closest centimetre. Weight was measured and body fat (%) and muscle mass (%) estimated using digital bio-impedance scales (BC-418, Tanita, USA). Waist and hip circumference were measured using a tape measure to the closest centimetre.

### Cardiopulmonary exercise testing (CPET)

A CPET was performed on a semi-recumbent cycle ergometer (Ergoselect1200, Ergoline, Germany) using a ramp protocol [12]. The test was terminated if the participant (i) reached 85% of their age-predicted (220-age) maximum heart rate (ii) experienced limiting symptoms or (iii) developed arrhythmia, hypotension (systolic blood pressure (BP) drop of >10mmHg despite increased workload) or (iv) an excessive blood pressure rise during the test (to >250 systolic or>115 diastolic mmHg).

Expired gases were analysed breath-by-breath, and heart rate and rhythm measured using a 6-lead ECG (Quark CPET, COSMED, Italy). Peak oxygen consumption (peak V·O_2_) was measured as the highest 30 second rolling average V·O_2_ value during exercise. Extrapolated V·O_2_max was calculated as peak V·O_2_ extrapolated to age-predicted maximum HR. Peak HR, oxygen uptake efficiency slope (OUES), anaerobic threshold (AT, if achieved), ventilatory equivalent of carbon dioxide (V·E:V·CO_2_) and ratio of oxygen uptake to work rate (V·O_2_/Work Rate) were all measured. Maximum V·O_2_ was predicted for men and women using the equations of Wasserman and Whipp [12]. Anaerobic Threshold was determined by both ventilatory equivalent and V-slope methods and is also presented as a percentage of predicted V·O_2_max. Respiratory exchange ratio (RER) was calculated as V·CO_2_/V·O_2._

Tests were conducted with continuous monitoring of the oxygen saturation of peripheral arterial blood (SpO_2_, finger or forehead probe) and BP was measured using a motion insensitive device (Tango M2, SunTech Medical, USA) every 2-3 minutes throughout exercise and recovery. Participants were asked to score both breathlessness (dyspnoea) and leg fatigue using the Borg CR10 scale on termination of exercise. Capillary lactate was measured in blood sampled from the fingertip prior to exercise and at peak effort using a Point-of-Care lactate analyser (Nova StatStrip Xpress, Nova Biomedical, UK).

### Lung function tests

Spirometry was performed in accordance with European Respiratory Society guidance [13] using an Easy-On-PC TrueFlow Spirometer (NDD Medical Technologies, France). Forced expiratory volume in one second (FEV_1_; % of predicted), forced vital capacity (FVC; % of predicted) and FEV_1_/FVC ratio are presented based on equations recommended by the European Respiratory Society and European Community of Coal and Steel. FEV_1_, FVC and FEV_1_/FVC Z-scores were calculated using the Global Lung Function Initiative reference equations [13].

### Cardiovascular function

Brachial blood pressure and heart rate readings were measured in the left arm with an appropriately sized cuff using a MIT Elite Plus (Omron, The Netherlands) in a resting seated position. Clinic BP and HR were estimated as the average of the final two of three consecutive readings.

Participants with Long COVID underwent a standard transthoracic echocardiogram (EPIQ 7G, Philips, MA, USA). Left ventricular (LV) structure and systolic and diastolic function were assessed using 2D, 3D and Doppler echocardiography. Full details of the protocol and outcome measures is provided in the supplementary information file (1.2.1).

In Long COVID cases, pulse wave velocity (PWV) was measured in a semi supine position using a Vicorder device (Skidmore Medical, Germany) according to manufacturer guidelines. Three consecutive measurements were acquired (within 0.5m/s of each other) and averaged. Full protocol details are described in the supplementary information file (1.2.2).

### Autonomic function

Heart rate recovery (HRR) was calculated at 1-minute and 2-minutes post-exercise. The 30 second rolling average HR centred around 60 and 120 seconds of the recovery phase was subtracted from the peak HR measured during exercise. In Long COVID cases, the change in BP and HR following a lying to standing manoeuvre were assessed using a brachial BP cuff (MIT Elite Plus, Omron, The Netherlands) to test for orthostatic hypotension (BP drop of >20 mmHg) and postural orthostatic tachycardia syndrome (POTS) (HR increase of >30 bpm on standing) [14, 15]. In Long COVID cases, a 5-minute 12-lead ECG was recorded at rest in the supine position to assess heart rate variability (HRV). Time domain (RMSSD the root mean square successive NN differences,) and frequency domain (LF low frequency, HF high frequency, LF normalised, LFHF ratio of low frequency to high frequency) measures were derived following manufacturer algorithms (CardioPerfect HRV Module 1.6.7.1149, Welch Allyn, USA).

### Muscle function

In Long COVID cases, dominant hand-grip strength (kPa; maximum of three measurements, with brief pauses between each) was measured with a pneumatic bulb hand dynamometer (Baseline, 3B Scientific, Germany). Concentric and eccentric strength of the left knee extensors/flexors was assessed at 60°/s for 5 repetitions using a HUMAC NORM isokinetic dynamometer (CSMi, USA). Participants were seated in an upright position with the lever arm pad secured proximal to the medial malleolus. The rotational axis of the knee was placed in line with the dynamometer axis of rotation, and 0° was determined as 0° knee extension. Tests were performed within a range of motion between 0 and 90° (individualised to each participant). The peak torque was defined as the highest point of the torque curve in the best repetition (Nm) and was normalised to body weight (Nm/Kg) for knee extensor and flexor, respectively.

### Skeletal muscle oxidative capacity, microvascular post-occlusive reactive hyperaemia (PORH) and changes in tissue saturation index (TSI) with exercise

Continuous wave (CW) NIRS (Portamon, Artinis Medical Systems, Netherlands) was used to assess skeletal muscle oxidative capacity and microvascular PORH. Full device and protocol details are provided in the supplementary information file (1.3.1).

Tissue saturation index (TSI), estimated using spatially resolved spectroscopy, was measured continuously from the gastrocnemius throughout the CPET. Greater decreases in TSI during exercise represent failure of oxygen supply to keep up with demand [16].

Adipose tissue thickness (ATT, average of three measures) overlying the NIRS measurement site was measured using an ultrasound device (EPIQ 7; Philips, USA) fitted with a high frequency transducer (L12-5; Philips, USA).

#### Post-processing of Near Infrared Spectroscopy (NIRS) data

NIRS data were analysed in MATLAB R2022a (MathWorks Inc, USA) using custom written programs as previously described [17], with post-processing fully described in the supplementary information file (1.3.2).

### Physical activity

#### Self-report

Participants were asked to complete the Recent Physical Activity Questionnaire (RPAQ), which assesses physical activity in four domains (leisure, work, commuting and home) during the past month [18]. Summary variables including the time (minutes/day) spent sedentary, and in light, moderate and vigorous activities, were derived using the MRC RPAQ data processing guidelines [19].

#### Wrist-worn actigraphy

Participants were fitted with a wrist-worn actigraphy monitor (Actiwatch Spectrum Plus, Philips, Netherlands) on their non-dominant wrist for 7-days following their clinic visit. Activity counts from the device over each 30 second epoch were used to determine sedentary, sleep and wake intervals and derive average activity counts per minute during the day.

#### Haemoglobin status

In Long COVID cases, capillary haemoglobin (g/L) was measured in blood sampled from the fingertip using a Point-of-Care analyser prior to exercise (Haemoglobin Hb 801, HemoCue, Sweden).

### Sample size

Sample size calculations were performed using GPower 3.1.9.7 (alpha = 0.05 (two-tailed) and 80% power) assuming that the minimum clinically important difference for the primary outcomes (CPET, lung, muscle and vascular function) corresponded to an effect size of 0.9 SD, (representing a 10-20% difference in outcome measure) based on previous studies.[20, 21] We aimed to recruit cases and controls in an approximate proportion of 2:1 to enhance the power of the planned sub-study analysis (described in full on ClinicalTrials.gov NCT04914754). On this basis, we calculated that 32 and 16 participants would be required in the Long COVID and control group respectively (48 in total).

### Statistical methods

Statistical analysis was performed in STATA 17.0 (StataCorp LLC, USA). Categorical participant data are presented as frequency (%) and continuous data presented as mean±SD if normally distributed or median [interquartile range; IQR] if skewed. For simple comparisons of two groups a Pearson’s Chi^2^ test was used to compare categorical variables and an unpaired Student’s t test (normally distributed), or Wilcoxon Rank Sum (skewed distribution) were used for continuous variables.

Outcome measures were compared between Long COVID and HC participants using causal inference methods to calculate differences and potential outcome means (POMs). This method aims to estimate the differences between groups while controlling for confounding bias present in observational studies. POMs were estimated using an augmented inverse probability weighted (AIPW) estimator with linear outcome and logit treatment models. AIPW is a statistical approach that combines propensity-based inverse probability weighting (where the contribution of an individual’s data is weighted by the propensity score) and regression adjustment. This approach has similarities to binary 1:1 matching, but is advantageous in that the entire sample is used and statistical power preserved. Additionally, AIPW is considered ‘doubly robust’, in that only one of the inverse probability weighting or regression adjustment need be correctly specified to obtain an unbiased effect estimator.

Estimates were adjusted for potential confounders chosen *a priori* (age, sex, ethnicity and BMI) informed by a directed acyclic graph and are presented as POM (95% confidence intervals) and differences between groups for model 1(M1). Regression diagnostics were performed, and histograms of propensity scores examined for appropriate overlapping. Missing data were dealt with via listwise deletion, which is valid under the assumption of missing completely at random.

Where well-accepted normal ranges exist, additional measures of cardiac, muscle, vascular and autonomic function were only performed in Long COVID cases and were compared to reference normative ranges or cut-offs for normal values. Confidence intervals for the n(%) abnormal were estimated using the Wilson method, and p-values calculated using binomial probability testing [22].

As an exploratory analysis, functional CPET outcomes were additionally adjusted for possible mediators: skeletal muscle function (muscle oxidative capacity, (Model 2(M2)), lung function (FEV_1_ Z-score; Model 3(M3)), autonomic function (HRR 2-min; Model 4(M4)) and combined (M2+M3+M4; Model 5(M5)), to establish if differences in one or all of these domains could explain some of the differences observed in performance at CPET. The natural indirect effect (NIE) refers to the difference between no mediator in the model and the effect after the mediator has been controlled by regression. The 95% confidence intervals and p-values for each NIE were calculated using bootstrapping with 1000 samples and the Wald test, respectively.

The level of significance was set at p<0.05. No adjustment was made for multiple testing, and inference was made based on effect size, statistical significance and 95% confidence intervals.

## Results

### Participant characteristics

In total, 32 participants with Long COVID (10(31%) men, 44±12 years old) and 19 healthy controls (6(32%) men, 40±13 years old) were recruited (Figure 1). Cases had higher BMI, waist-to-hip ratio, body fat (%) and calf adipose tissue thickness than controls (Table 1). Amongst cases, 26(81%) of participants self-reported a pre-existing condition including: hypertension, asthma, type 2 diabetes mellitus and mental health conditions (supplementary Table S1).

**Figure 1.**
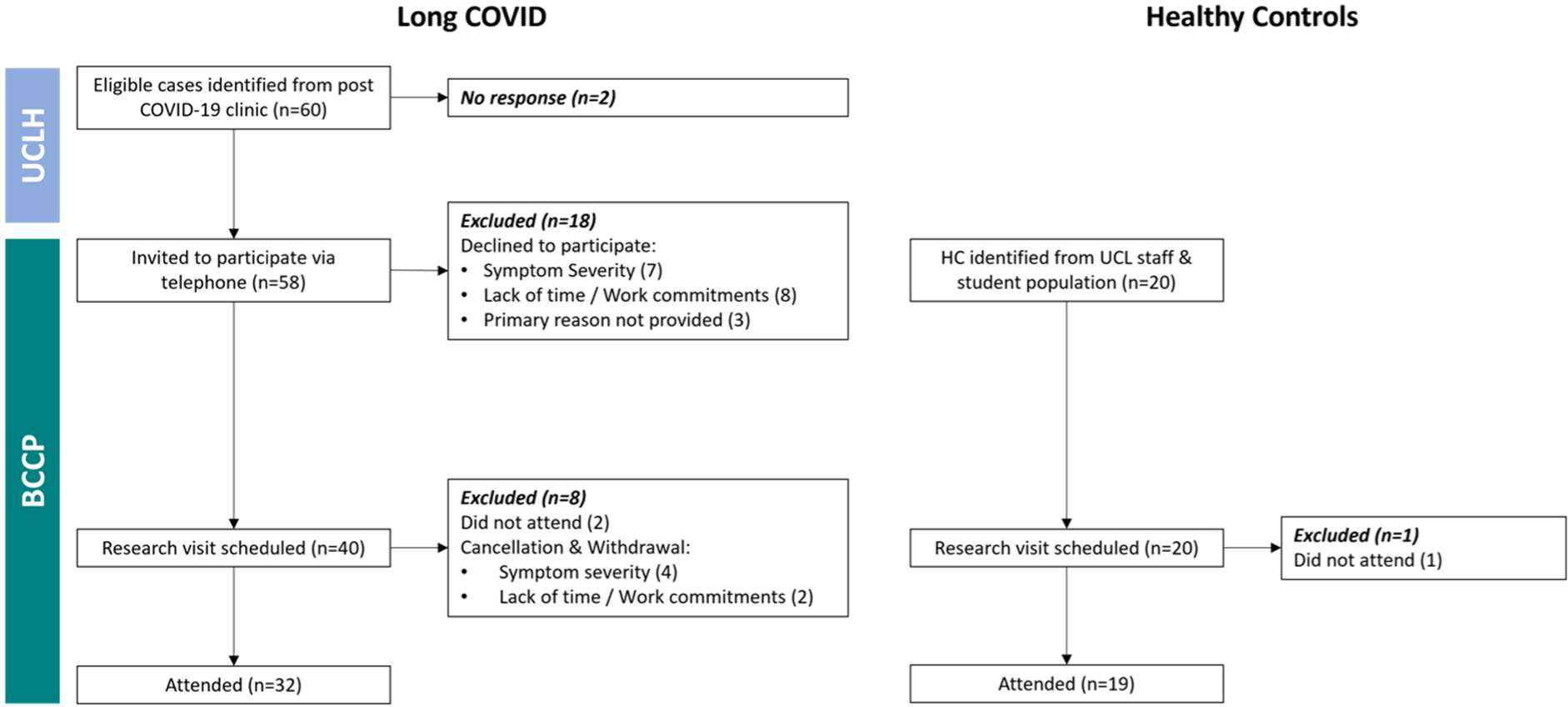
Study participant recruitment flowchart. Participants with Long COVID with self-reported exercise intolerance and fatigue were identified from the post COVID-19 clinic at University College London Hospital (UCLH). The study research team at Bloomsbury Centre for Clinical Phenotyping (BCCP) invited eligible individuals to attend a research visit. Healthy Control participants sex and age (5-year banding) were recruited from the staff and student population at UCL.

**Table 1.** Characteristics for 32 Long COVID cases versus 19 healthy controls. BMI (body mass index), SBP & DBP (resting systolic and diastolic blood pressure), HR (heart rate), ATT (Adipose Tissue Thickness).

|  | Mean±SD, median[IQR] or n(%) |  |  |
| --- | --- | --- | --- |
|  | Healthy Controls<br>(n=19) | Long COVID<br>(n=32) | p-value |
| Age [years] | 40±13 | 44±12 | 0.235 |
| Male Sex | 6(32%) | 10(31%) | 0.981 |
| Ethnicity |  |  |  |
| White European | 15(79%) | 20(63%) | 0.360 |
| Indian Asian | 3(16%) | 7(22%) |  |
| Black African Caribbean | 0(0%) | 4(12%) |  |
| Mixed | 1(5%) | 1(3%) |  |
| BMI [kg/m<sup>2</sup>] | 24.9±3.7 | 28.4±5.8 | 0.022 |
| Waist-to-Hip Ratio | 0.82[0.09] | 0.88[0.10] | 0.029 |
| Clinic SBP [mmHg] | 119±14 | 124±19 | 0.397 |
| Clinic DBP [mmHg] | 79±11 | 82±13 | 0.495 |
| Resting HR [bpm] | 73±12 | 74±13 | 0.877 |
| Current Smoker | 0(0%) | 2(6%) | 0.270 |
| No. of comorbidities | 0 | 2±2 | <0.001 |
| No. of Long COVID symptoms | 0 | 10±4 | <0.001 |
| Body Fat [%] | 28±7 (n=18) | 34±7 (n=28) | 0.008 |
| Estimated Muscle Mass [%] | 69±7 (n=18) | 62±7 (n=28) | 0.002 |
| Calf ATT [cm] | 0.61±0.16 (n=17) | 0.83±0.34 (n=25) | 0.016 |

#### Acute SARS-CoV-2 severity and treatment

Thirty cases had been managed at home during their acute SARS-CoV-2 infection, and 2 had required hospitalisation, one received dexamethasone and tocilizumab and the other received dexamethasone and remdesivir.

#### Long COVID symptomatology

Cases were investigated in our study, on average, 14±6 months post-acute SARS-CoV-2 infection. On average participants with Long COVID reported 10±4 symptoms, and all were experiencing fatigue and exercise intolerance (supplementary Figure S1).

### Long COVID versus healthy control participants and normative values

#### Exercise performance and cardiopulmonary fitness

Four participants with Long COVID had a contraindication to exercise testing on the day of their study visit and were excluded. A further two participants with Long COVID were unable to exercise beyond the warm-up phase and were excluded from subsequent analyses. Only 8(29%) cases achieved 85% of their age-predicted maximum heart rate compared with 14(74%) of controls (p=0.001). Of the 20 remaining cases, 15 experienced limiting symptoms, 4 had a hypotensive response (systolic BP drop of >10mmHg with increased workload) in the absence of ischaemic ECG changes, and 1 had an exaggerated exertional blood pressure response (>250/115 mmHg).

After adjustment for age, sex, ethnicity and BMI, cases achieved a lower exertional V·O_2_peak and had lower extrapolated V·O_2_max than controls (Table 2). OUES, anaerobic threshold, peak HR, O_2_ pulse and ratio of oxygen uptake to work rate were also lower, and VE/V·CO_2_ slope higher (Table 2). A respiratory exchange ratio (RER) of ≥1.1 was measured in 22(85%) of cases and 17(90%) HC. Borg CR10 scores were lower in HC than cases; (5±1 versus 6±2 for breathlessness and 6±3 versus 7±2 for leg fatigue). Sensitivity analysis directly comparing 18 HC and 18 cases, unadjusted for confounders, showed similar trends (supplementary Table S2).

**Table 2.** Cardiopulmonary Exercise Test confounder adjusted (age, sex, ethnicity, & BMI) results for Healthy Controls versus Long COVID cases. V O_2_ (Oxygen Uptake), AT (Anaerobic Threshold), V E/V CO_2_ (Ventilation/Carbon Dioxide Production), OUES (Oxygen Uptake Efficiency Slope), V O_2_WR (Oxygen Uptake Work Rate), RER (Respiratory Exchange Ratio).

|  |  | Potential Outcome Means(95%CI)<br>Adj. for age, sex, ethnicity & BMI |  | The difference in Long COVID vs HC<br>M1: Adj. for age, sex, ethnicity & BMI |  |
| --- | --- | --- | --- | --- | --- |
|  | n | Healthy Controls | Long COVID | Δ (95%CI) | p-value |
| Cardiopulmonary Exercise Test |  |  |  |  |  |
| Peak $\dot{V}O_2$ [ml/kg/min] during exercise | 45 | 23.3(21.4,25.2) | 18.3(16.6,19.7) | -5.0(-7.1,-3.0) | <0.001 |
| Extrapolated $\dot{V}O_2$ max [ml/kg/min] | 45 | 33.0(30.7,35.3) | 30.5(28.1,32.8) | -2.5(-5.2,0.2) | 0.064 |
| OUES [ml/min] | 45 | 2175.8(1978.3,2373.3) | 1847.4(1678.5,2016.4) | -328.3(-534.4,-122.3) | 0.002 |
| $\dot{V}O_2$ at AT [ml/Kg/min] | 44 | 15.6(14.4,17.2) | 13.2(12.2,14.3) | -2.4(-3.7,-1.1) | <0.001 |
| VAT [% of predicted] | 44 | 56.7(53.3,60.0) | 47.8(44.1,51.6) | -8.8(-13.3,-4.4) | <0.001 |
| $\dot{V}E/\dot{V}CO_2$ Slope | 45 | 25.7(24.5,26.8) | 28.0(26.9,29.2) | 2.4(0.9, 3.8) | 0.001 |
| Peak HR [min<sup>-1</sup>] | 45 | 146(143,149) | 130(119,140) | -16(-27,-5) | 0.003 |
| Peak HR [% predicted] | 45 | 82(81,84) | 73(68,79) | -9(-15,-3) | 0.002 |
| Highest $\dot{V}O_2$/HR (O<sub>2</sub> pulse) [ml/beat] | 45 | 12.0(10.8,13.1) | 10.5(9.6,11.3) | -1.5(-2.5,-0.5) | 0.002 |
| $\dot{V}O_2$/WR [ml/min/W] | 45 | 9.3(8.9,9.8) | 8.7(8.2,9.1) | -0.7(-1.3,-0.1) | 0.027 |
| Peak Power [W] | 45 | 170.5(154.5,186.4) | 128.2(112.9,143.4) | -42.3(-56.6,-28.0) | <0.001 |
| Peak Ventilation [L/min] | 45 | 58(52,64) | 48(41,54) | -11(-18,-4) | 0.001 |
| Peak Breathing Reserve [%] | 45 | 57(52,63) | 59(53,66) | 2(-6,11) | 0.607 |
| Peak RER | 45 | 1.18(1.15,1.21) | 1.11(1.06,1.17) | -0.07(-0.13,-0.01) | 0.024 |
| No. of participants RER≥1.1 |  | 22(85%) | 17(90%) | - | - |
| Lactate [Δmmol/L] | 36 | 4.5(3.5,5.5) | 2.6(1.5,3.7) | -1.9(-3.2,-0.6) | 0.003 |
| Borg CR10 Dyspnoea | 44 | 5(5,6) | 6(5,6) | 0(-1,1) | 0.500 |
| Borg CR10 Leg Fatigue | 45 | 6(4,7) | 7(6,8) | 1(-0,3) | 0.139 |

Of the participants with Long COVID, 32% fell below the normal cutoff for extrapolated V·O_2_max, 31% for OUES and 46% for V·O_2_/WR. 14% met criteria for V·E/V·CO_2_ abnormality (slope >30), but we did not observe any individuals who were below normal limits for breathing reserve (<15L).

#### Lung function

FEV1 and FVC (percentage of predicted and Z-scores) were lower in cases than controls (Table 4). However, means all fell within normal ranges (above LLN) and 69% had normal spirometry as defined by FEV_1_, FVC and FEV_1_/FVC Z-score > -1.64.

#### Cardiac structure and function

We did not find strong evidence of systolic or diastolic cardiac dysfunction in cases assessed at rest by echocardiography. The prevalence of abnormal LV relaxation (e’) in 19-25% of Long COVID individuals was higher than anticipated, although none met clinical criteria defining diastolic dysfunction. All participants fell within normal limits for LVEF measured by 3D echocardiography, a method that avoids geometric assumptions (Table 3).

**Table 3.**
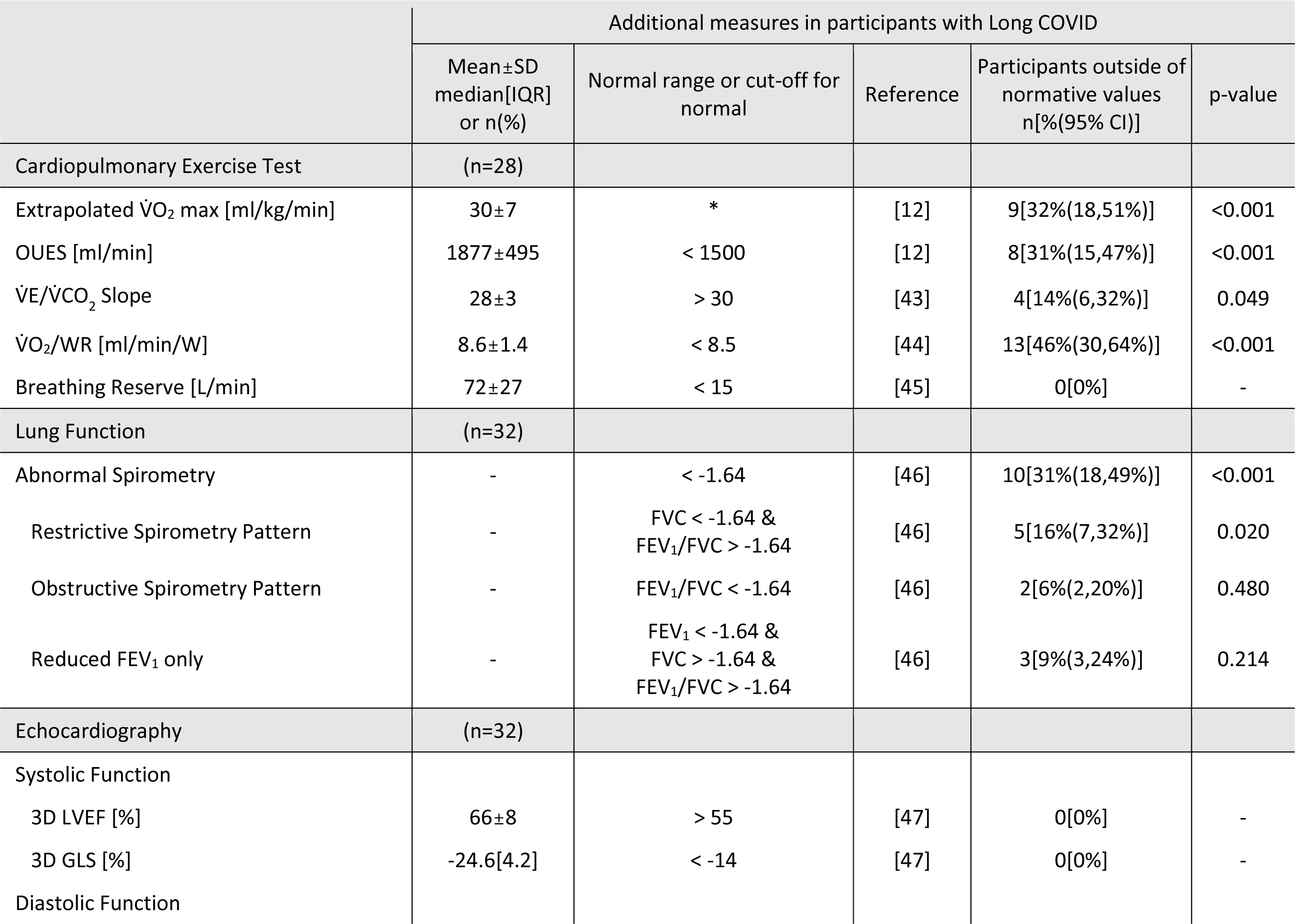
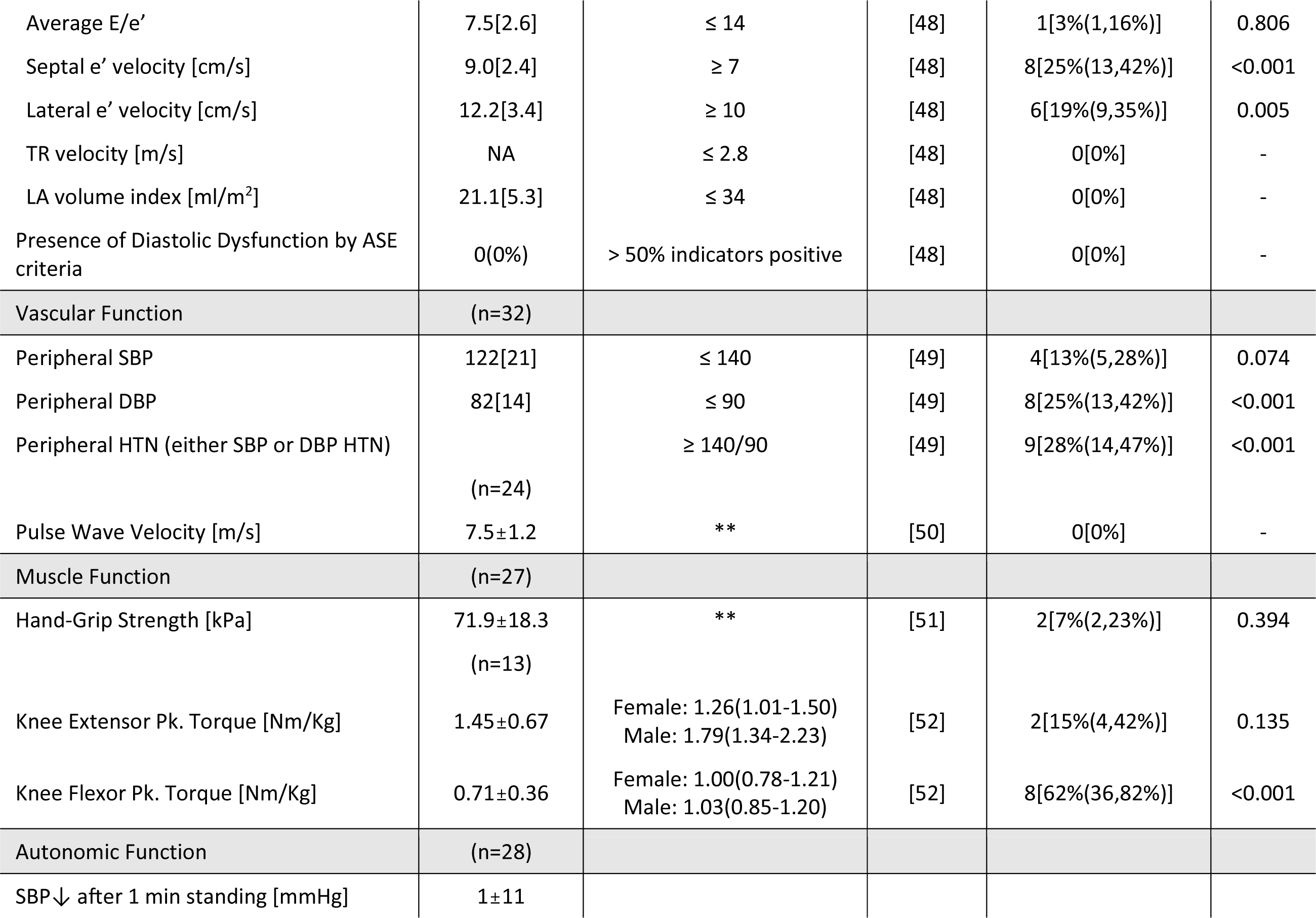
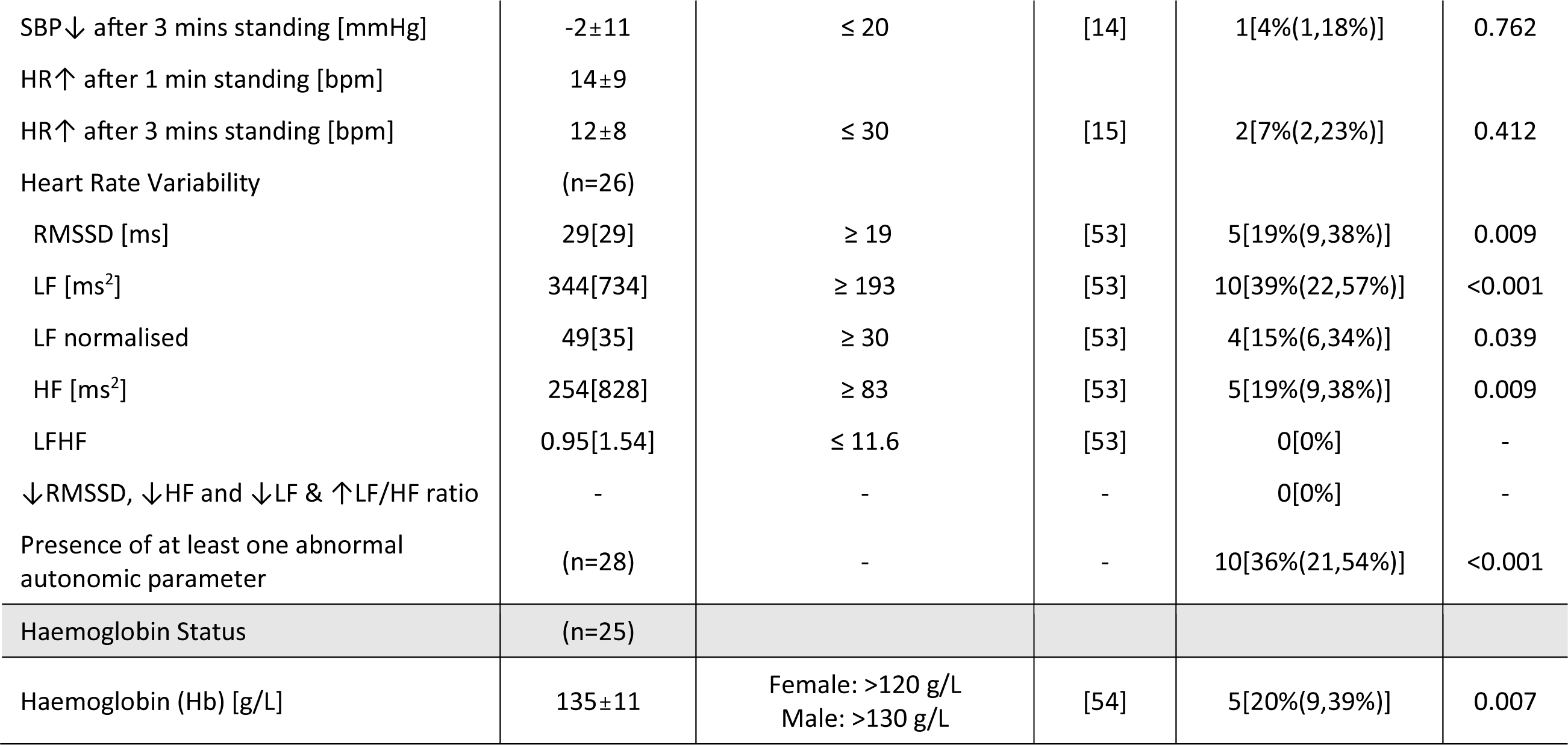
Additional measures of cardiac, vascular, muscle and autonomic function and haemoglobin status for participants with Long COVID. *Normative values were calculated using sex specific equations. **Normative values were stratified by sex and age decade. Details are provided in full in references provided. V O_2_ (Oxygen Uptake), V E/V CO_2_ (Ventilation/Carbon Dioxide Production), OUES (Oxygen Uptake Efficiency Slope), V O_2_WR (Oxygen Uptake Work Rate), FEV_1_ (Forced Expiratory Volume in the 1^st^ second), FVC (Forced Vital Capacity), FEV_1_/FVC ratio, LVEF (Left Ventricular Ejection Fraction), GLS (Global Longitudinal Strain), TR (Tricuspid Regurgitation), LA (Left Atria), ASE (American Society of Echocardiography), Pk (Peak), SBP (Systolic Blood Pressure), DBP (Diastolic Blood Pressure), HR (Heart Rate), RMSSD (Root Mean Square of Successive Differences between normal beats), LF (Low Frequency), HF (High Frequency), LFHF (Low Frequency High Frequency ratio).

**Table 4.**
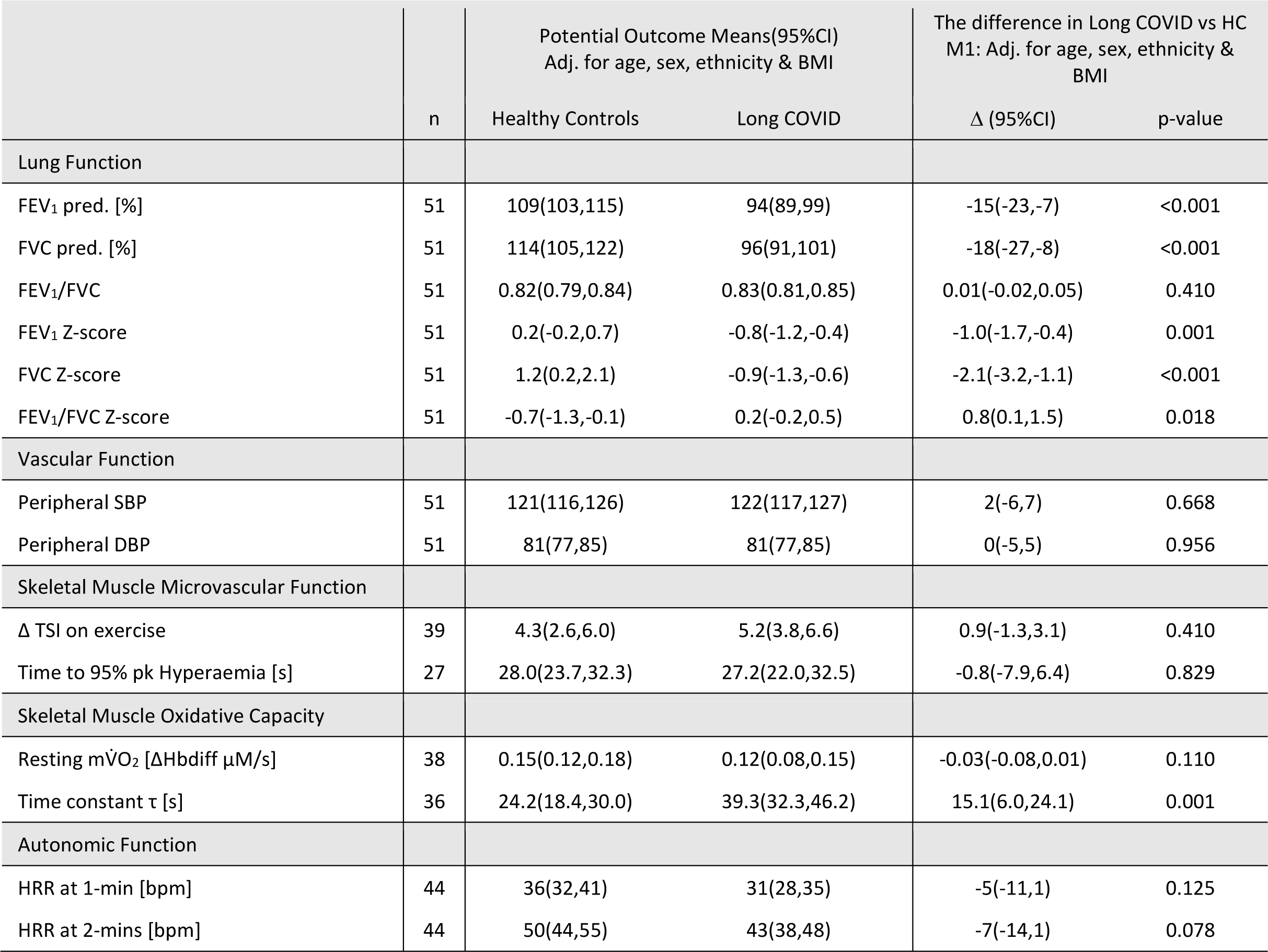
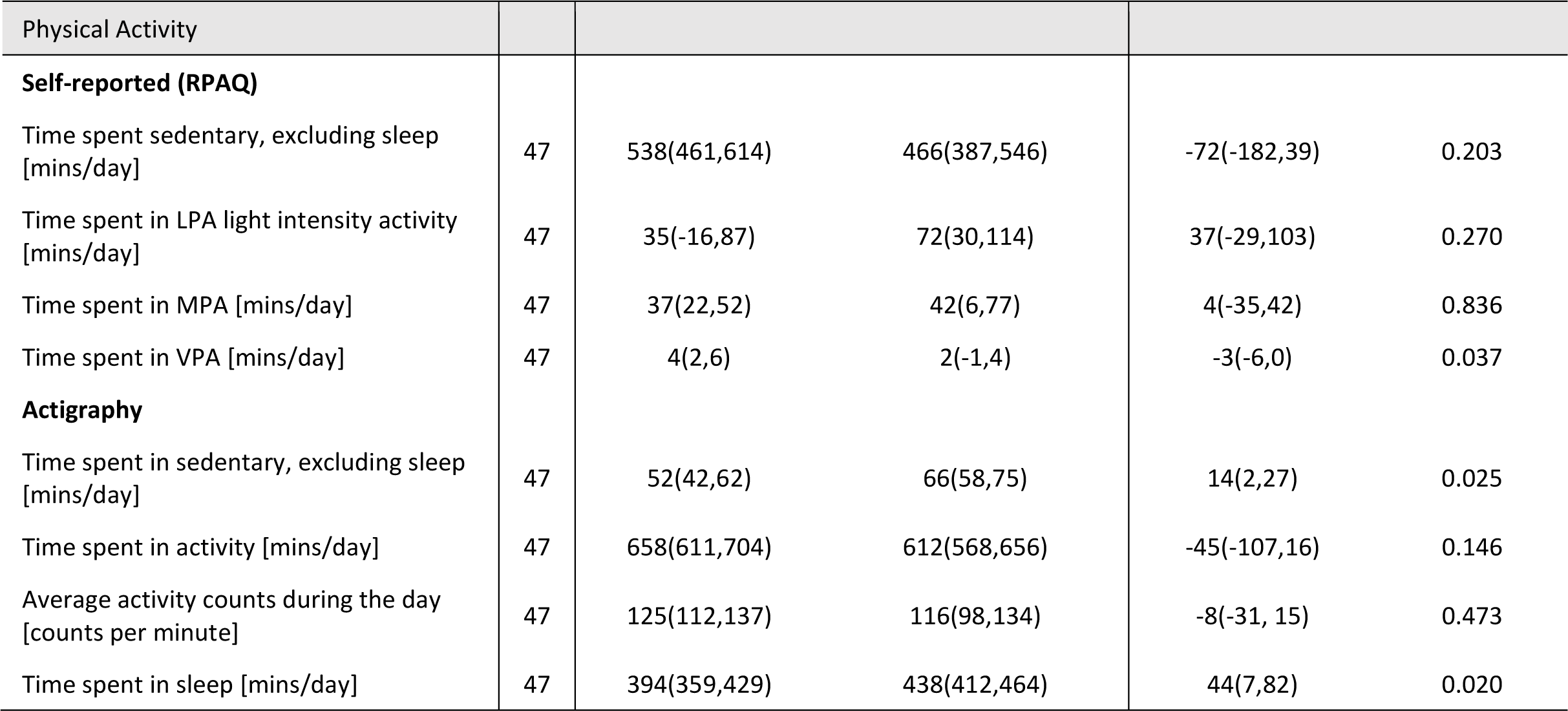
Lung Function (Spirometry), Vascular Function (Resting Blood Pressure), Skeletal Muscle Function (Near Infrared Spectroscopy), Microvascular Function (Near Infrared Spectroscopy), Autonomic Function (Heart Rate Recovery) and Physical Activity (self-reported and actigraphy) confounder adjusted (Age, Sex, Ethnicity, & BMI) results for Healthy Control participants versus Long COVID cases. FEV_1_ (Forced Expiratory Volume in the 1^st^ second), FVC (Forced Vital Capacity), FEV_1_/FVC ratio, SBP (Systolic Blood Pressure), DBP (Diastolic Blood Pressure), TSI (Tissue Saturation Index), mV O_2_ (Muscle Oxygen Uptake), HRR (Heart Rate Recovery), RPAQ (Recent Physical Activity Questionnaire), LPA (light physical activity), MPA (moderate physical activity), VPA (vigorous physical activity).

#### Skeletal muscle function

A total of six NIRS recordings were rejected: five due to apparent failure of arterial occlusion, and one due to the participant not fully recovering from the exercise after 3-minutes of transient occlusions (data did not fit to the mono-exponential curve and plateau not achieved). Resting muscle oxygen consumption (musV·O_2_) was similar but the time constant for the recovery of muscle V·O_2_ was longer in cases than controls (Table 4). Complete arterial occlusions for the PORH measure were on average 240±60 seconds. There were no differences in time to 95% peak hyperaemia and change in TSI during exercise (Table 4). Hand- grip strength was 71.9±18.3 kPa and knee extensor/flexor peak torque was 1.45±0.67 Nm/Kg and 0.71±0.36 Nm/Kg, respectively (Table 3). Muscle strength measures were below normal limits in 7% of cases for hand-grip strength, 15% for knee extensor, and 62% for knee flexor.

#### Autonomic function

Despite achieving a lower peak HR, HR change after 1 minute of recovery was similar between cases and controls. However, HR change after 2 minutes of recovery was less in cases (Table 4). One individual met the criteria for orthostatic hypotension and 2(7%) individuals met the criteria for POTS. At least one resting HRV parameter fell outside normative ranges in 39% of cases (Table 3). Additional HRV parameters are summarised in the supplementary information file (Table S3).

#### Physical activity (PA)

Differences were not observed between groups in self-reported RPAQ time spent in sedentary, light and moderate PA. However, cases reported spending less time in vigorous activity than controls (Table 4). PA assessed via actigraphy showed more time sedentary 66(58,75) vs (52(42,62) mins, p=0.025) and sleeping (438(412,464) vs 394(359,429) mins, p=0.020) in cases than controls. However, differences were not observed in the time spent in activity (612(568,656) vs 658(611,704) mins, p=0.473) and average number of activity counts per minute (116(98,134) vs 125(112,137) mins, p=0.146) between groups (Table 4).

#### Haemoglobin status

5(20%) cases fell below the normal cutoffs for haemoglobin levels (Table 3), three of which were included in CPET analyses.

### Mediation analysis

To explore whether differences in skeletal muscle, lung, autonomic and cardiac function (separately and combined) might explain the differences in exercise performance observed between cases and controls, we performed an exploratory analysis in which we further adjusted our models for the following: time constant (M2), FEV_1_ Z-score (M3), HRR 2-mins (M4), and M2+M3+M4 combined (M5). Model 1-5 estimations (95% confidence intervals) for key CPET outcomes are illustrated as forest plots in Figure 2.

**Figure 2.**
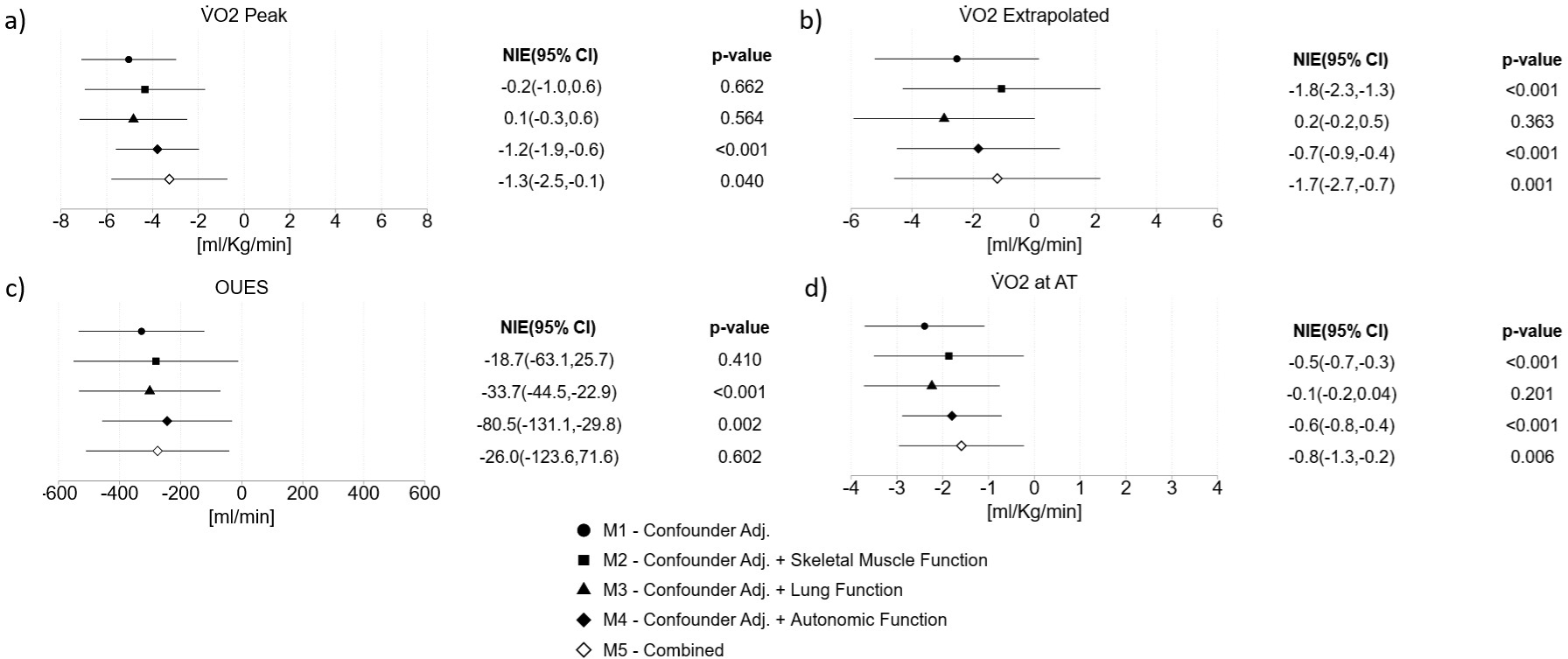
Forest plots for the mediation analysis of cardiopulmonary exercise test outcomes. The natural indirect effect (NIE) refers to the difference between no mediator in the model and the effect after the mediator has been controlled by regression. The 95% confidence intervals and p-values relate to each NIE. Model 1 (M1): confounder adjusted for age, sex, ethnicity and body mass index (BMI); Model 2 (M2): confounder adjusted + adjusted for skeletal muscle function (time constant); Model 3 (M3): confounder adjusted + adjusted for lung function (FEV_1_ Z-score); Model 4 (M4): confounder adjusted + adjusted for autonomic function (heart rate recovery (HRR) at 2-mins); and Model 5 (M5): confounder adjusted + adjusted for time constant, FEV_1_ Z-score and HRR at 2-mins (combined). V O_2_ (Oxygen Uptake), AT (Anaerobic Threshold), OUES (Oxygen Uptake Efficiency Slope).

The differences in extrapolated V·O_2_max between cases and controls were attenuated from -2.9 to -1.1 (p<0.001) by the time constant (M2) and -2.5 to -1.8 (p<0.001) by HRR 2-mins (M4). Evidence of partial mediation was observed in each of the investigated systems: for the time constant (M2), this included V·O_2_ at AT; for FEV_1_ Z-score (M3), this included OUES and V·O_2_ at AT and for HRR at 2-mins (M4), this included OUES.

## Discussion

Individuals experiencing symptoms of COVID beyond 12 weeks post-infection have reduced skeletal muscle oxidative capacity, lung function and ANS function compared to age, sex and BMI-matched healthy control participants. Compared to normal clinical ranges, ∼30% of participants with Long COVID fell outside the normal range for spirometry and 36% for ANS function. However, CPET indices suggest exercise performance was limited by peripheral factors, rather than lung or circulatory dysfunction. Through exploratory mediation analyses, we did not find strong evidence that any one physiological deficit fully explained the severely impaired exercise performance in Long COVID; this points to a multi-system dysfunction contributing to exercise impairment. It is important to recognise that our study sample was small for mediation analysis and we interpret these results with caution.

The observed reduction in exercise performance in Long COVID is aligned with prior studies [4, 5, 8, 23–32]. We present maximal and sub-maximal indices of cardiopulmonary fitness, all indicating impairments in performance and cardiopulmonary fitness. O_2_ pulse was lower in cases than controls in line with prior findings [33, 34]. A low O_2_ pulse can indicate impaired augmentation of stroke volume or impaired peripheral oxygen extraction during exercise. The reduced AT and V·O_2_/WR ratio in Long COVID cases point towards a peripheral limitation. This is in line with findings by Singh *et al* who reported that exercise capacity beyond 75% of peak V·O_2_ was limited by a failure to increase peripheral oxygen extraction [5].

Our finding that individuals with Long COVID had poorer skeletal muscle oxidative capacity further supports a peripheral skeletal muscle impairment in the presence of Long COVID. This is concordant with a study by Colosio et al who performed high resolution respirometry on skeletal muscle biopsies and non-invasive NIRS measurements, similar to our NIRS measurements, in people with Long COVID and healthy controls [6]. Together, these findings provide evidence for a deficit in skeletal muscle mitochondrial bioenergetics in Long COVID. Our larger sample size, compared to Colosio et al, allowed us to perform additional statistical analyses exploring the role of oxidative capacity in mediating the effect of Long COVID on exercise performance. When we included oxidative capacity (τ) as a mediator in statistical models, we observed that the differences observed in extrapolated V·O_2_max and V·O_2_ at AT between cases and controls were partially attenuated. Partial mediation was also observed when we included the ANS system and lung function measures into the model, therefore, we cannot exclude the possibility of multi-system involvement in the exercise performance impairment observed in Long COVID participants. Our study sample size is small for this type of analysis which limits our power to perform multivariable adjustments, and this could have impacted our results. Larger studies are necessary to confirm these effects.

Support for a skeletal muscle deficit more generally was also evidenced by the reduced knee extension/flexion force in 62% of Long COVID cases enrolled here. This may be explained by the reduced lean mass in our study group, as was described by Ramirez-Velez and colleagues [35]. Only 7% of individuals with Long COVID had a hand-grip strength outside the normal range [35], which may suggest a detraining effect on the lower limbs in line with the increase in sedentary time that we observed objectively. However, we also report similar self-reported and objectively measured levels of physical activity between groups which would argue against detraining. A major limitation of this study, and many other Long COVID studies, is that we cannot be certain whether the deficits described existed prior to COVID-19 infection or are the result of infection. Studies where antecedent measurements have been performed would be extremely useful in understanding the development of pathophysiology in Long COVID.

There was no strong evidence of resting cardiac structural or functional abnormalities or evidence of increased stiffening of the arterial system in our Long COVID cases. Time to 95% PORH measured using NIRS was similar in cases and controls. This suggests reactivity in the microcirculation, an indicator of endothelial function, is not impaired in Long COVID [16]. In 4 Long COVID participants resting blood pressure was above the cut-off for both systolic and diastolic hypertension (>140/90mmHg). All 4 participants reported a diagnosis of hypertension prior to COVID-19 infection and were being treated with anti-hypertensive agents, none were treated with a β-blocker.

### Autonomic function

There was heterogeneity across measures of autonomic function in Long COVID cases. On average, HR recovered more rapidly in controls than in Long COVID cases, in line with previous work [4, 7, 36–38]. During the lying to standing manoeuvre, one individual met the clinical criteria for orthostatic hypotension and a further two met the criteria for POTS [9]. We also observed exaggerated heart rate increases on standing (>20bpm) in a further 7 individuals, suggesting possible sub-clinical abnormalities. A limitation is that HR and BP measurements were not continuous, meaning that we were only able to assess single measurement differences, increasing the possibility of measurement errors. No Long COVID participants fell outside of normal limits for all three key HRV variables measured (LF, HF and RMSSD), however, 39% fell below normal limits for the LF variable and 18% below normal limits for the RMSSD and HF variables, a finding in line with HRV alterations previously described in Long COVID case-control studies [39, 40].

## Strengths and limitations

Causal inferences are limited in our study by small sample size and cross-sectional design. We cannot be certain whether observed pathophysiology in Long COVID participants was the cause or consequence of their symptoms, or secondary to their pre-existing (non-COVID) states. Likewise, we cannot confirm whether differences in body composition were due to Long COVID-related inactivity or a secondary (or pre-existing) low grade metabolic syndrome that would predispose to poorer oxidative capacity in muscle. We performed additional measures of cardiac, autonomic, vascular and skeletal muscle function in participants with Long COVID only and used reference normative values to identify those who fell outside of normal limits. However, we cannot rule out the fact that these sub-clinical values might not have also been observed in the general population at the same frequency. We used NIRS to measure oxidative capacity, and the PORH response to ischaemia as a measure of microvascular function [41]. There are some limitations to NIRS, discussed in detail elsewhere [42]. The gold-standard non-invasive method for assessing oxidative capacity is to directly measure PCr recovery using ^31^P-MRS, confirmation of these findings via this method would be useful.

One limitation is that CPET was terminated at 85% of predicted maximum HR. This exercise protocol was established in consideration of safety concerns during the early stages of the pandemic when the study was designed and in accordance with the restrictions imposed by the research ethics committee. Exercise was limited by symptoms in 58% of Long COVID cases providing a measured maximal V·O_2_ in those individuals, and demonstrated reduced cardiopulmonary fitness. Furthermore, a RER of ≥1.1 was measured in 85% of cases and 90% of controls indicating that the majority of individuals were nearing peak effort.

## Conclusions

We have identified a limitation in peripheral oxygen uptake with normal local vascular supply in the presence of Long COVID, suggesting pathophysiology of mitochondrial oxygen uptake and utilisation. Understanding the exact mechanism of the myocellular defect is important to identify therapeutic targets. We also observed some evidence for lung and autonomic dysfunction highlighting that exercise intolerance in Long COVID may not be attributed to impairment in a single physiological system, but rather the result of an accumulation of multi- system, often sub-clinical, dysfunction and their interplay.

## Data Availability Declaration

Due to the sensitive nature of the data collected for this study, data cannot be made publicly available, but requests to access the dataset from qualified researchers trained in human subject confidentiality protocols may be sent to Dr Siana Jones at the MRC Unit for Lifelong Health and Ageing at UCL.

## Acknowledgements

We are extremely grateful to all the people who took part in this study and to the past and present members of the research team at BCCP who helped to collect the data.

