## Supplementary Information File for "Mechanisms underlying exercise intolerance in Long COVID: an accumulation of multi-system dysfunction"

### Methods

#### 1.1 Exclusion criteria

Individuals were excluded if they were considered a vulnerable adult, were unable to consent, had a severe or terminal illness, were pregnant, had a known contra-indication to exercise testing, or were using corticosteroids (e.g. any maintenance dose of oral steroid, 4 or more courses of oral steroid in previous 12 months, but not including prescription of dexamethasone during the acute episode of COVID-19, as Long as it had been discontinued).

#### 1.2 Cardiovascular function

##### *1.2.1 Echocardiography*

Participants with Long COVID underwent transthoracic echocardiography in the left lateral decubitus position using an EPIQ 7G Ultrasound System equipped with a X5-1 transducer (Philips, MA, USA). Echocardiographic measures were obtained from the parasternal long-axis (PLAX); parasternal short-axis; and apical four-, two- and five-chamber views and were performed by three sonographers in accordance with the American Society of Echocardiography guidelines [1].

Left ventricular (LV) systolic and diastolic function were assessed using TOMTEC software (version TAA2.40, TOMTEC Imaging Systems GmbH) by a single experienced echocardiographer who was blinded to the objectives of the project and was not present during data capture. Standard 2D- and Doppler-echocardiographic measurements were performed offline following the ASE/EACVI guidelines [2, 3]. Left ventricular (LV) volumes and LV ejection fraction (LVEF) were calculated using the modified biplane Simpson's rule method and using the 4D analysis package. Maximum left atrial volume indexed (LAVi) to body surface area was calculated by the biplane method of discs at end-systole with LA enlargement defined as LAVi >34 mL/m$^2$. LV diastolic function was evaluated in accordance with the current ASE/EACVI guidelines. This included mitral inflow diastolic filling velocities [(E-wave and A-wave), *E*/*A* ratio, and deceleration time (DT)], tissue-Doppler analysis of lateral and septal mitral annular motion (e′, a′, and s′) from which averaged *E*/*e*′ ratio was calculated, and

Doppler derived-pulmonary artery systolic pressure (PASP) estimated from the peak tricuspid regurgitation (TR) velocity jet. As per ASE guidelines, diastolic dysfunction was identified as greater than 50% of the following parameters meeting cutoff values: septal e′ < 7 cm/sec, lateral e′ < 10 cm/sec), average E/e′ ratio > 14, LA maximum volume index > 34 mL/$m^2$, and peak TR velocity > 2.8 m/sec [3].

*1.2.2 Pulse Wave Velocity (PWV)*

The neck cuff was positioned over the left carotid pulse and the leg cuff was positioned as high as possible on the left thigh. The distance from the suprasternal notch and the neck (proximal) and thigh (distal) cuffs were measured using a measuring tape, respectively. The distal distance was entered into the Vicorder software prior to data acquisition. Approximately 10 sequential waveforms of the carotid and femoral arteries were reviewed by eye prior to data capture.

1.3 Muscle function

*1.3.1 Near Infrared Spectroscopy (NIRS)*

This device uses continuous wave (CW) NIRS to measure relative changes in oxy- and deoxy-haemoglobin (Hb). With the participant positioned supine, the NIRS device was placed over the left lateral gastrocnemius. The device was secured using micropore tape and covered using a black neoprene sleeve to prevent ambient light interference. A leg cuff was placed above the left knee, proximal to the measurement site. The device was set to sample at 10Hz. After stabilisation of the trace, an arterial occlusion was imposed by rapidly inflating the cuff (a rapid cuff inflation system, Hokanson, United States) to >250 mmHg for 30 s. Local skeletal muscle oxygen consumption (mus$\dot{V}O_2$) was estimated from the change in oxy- and deoxy-Hb [4]. Following a 5-minute plantar flexion resistance band exercise protocol (at a rate of 30 plantar flexions a minute), short transient arterial occlusions lasting 5-8 seconds were applied over 3 minutes to track recovery of mus$\dot{V}O_2$ and measure the recovery time constant ($\tau$), an estimate of oxidative capacity [5-7]. Longer values of $\tau$ represent poorer skeletal muscle oxidative capacity [5].

After a further recovery period, the leg-cuff was inflated again to >250 mmHg for at least 2 minutes. We aimed to have a 5-minute arterial occlusion in all participants, however, this measure was poorly tolerated in our study. Based on prior work demonstrating an association between flow and vessel dilatation as measured by flow mediated dilatation (FMD) with 1.5 minutes arterial occlusion, we set a threshold of 2 minutes for inclusion in the results [8]. The cuff was then released, and the PORH response was recorded for a minimum of 3 minutes.

*1.3.2 Post-processing of Near Infrared Spectroscopy (NIRS) data*

Slope values were multiplied by -1 for ease of interpretation; higher values represent higher muscle oxygen consumption. Post-exercise mus$\dot{V}O_2$ was estimated from the $Hb_{diff}$ signal during each of the transient arterial occlusions performed after exercise. The time constant of recovery, τ, was derived from a mono-exponential curve fitted to the post-exercise mus$\dot{V}O_2$ measures. Time to 95% and 100% peak post occlusive reactive hyperaemia (PORH) were calculated using methods previously described as an estimate of microvascular function [9-11]. The TSI signal during CPET was measured at (I) baseline, (II) minimum observed during exercise and (III) end of exercise, as previously described [11], and differences calculated.

Analysis of NIRS data was performed in MATLAB R2022a (MathWorks Inc, USA) using custom written programs and performed by a single observer. Signals were rejected if there was evidence that complete arterial occlusion had not been achieved (pulsatility in the oxy-Hb and deoxy-Hb signals and/or lack of reciprocal changes in oxy-Hb and deoxy-Hb signals). Resting mus$\dot{V}O_2$ was estimated by fitting a slope to the NIRS signal calculated from difference between oxy-Hb and deoxy-Hb (Hbdiff). A 10 second portion of this signal close to the start of the resting, 30 second arterial occlusion was selected [12]. Although these slopes were negative, we multiplied values by -1 to present these results as positive values for ease of interpretation; higher values represent higher muscle oxygen consumption.

Post-exercise mus$\dot{V}O_2$ was estimated from the Hbdiff signal during each of the transient arterial occlusions performed after exercise. The signals were visualised by eye and a 4-second period selected for each slope. A time constant, τ, was derived from a mono-exponential curve fitted to the mus$\dot{V}O_2$ measures captured post-exercise. Longer values of τ are considered to represent poorer skeletal muscle oxidative capacity [5].

The TSI signal during CPET was visualised and 5 second measurement periods were selected by eye where the signal was: stable at rest (baseline), had reached an initial peak at the onset of exercise (increment), at the minimum observed during exercise and at the end of exercise as previously described [11]. The difference between TSI at baseline and at the initial increment was calculated as the increment minus the baseline value. The TSI change from baseline to end of exercise was calculated as the baseline value minus the value at the end of exercise. The TSI change from the increment to the minimum was calculated as the increment value minus the minimum value. Greater drops in TSI during exercise are thought to represent failure of oxygen supply to keep up with demand [13].

Time to 95% and 100% peak post occlusive reactive hyperaemia (PORH) were calculated using methods previously described as an estimate of microvascular function [9, 10]. In brief, traces were inspected by eye and the point at which the cuff was released was identified from the oxy-Hb trace as the first upward inflection point. The peak response was identified as the maximum value captured within 2 minutes from cuff release. The change in oxy-Hb from the start to the end of the PORH (ΔOxyHb) was calculated as the peak value minus the value for cuff release. The recovery rate was calculated as 1 divided by the time to 100% peak PORH and is presented in seconds [11].

Results

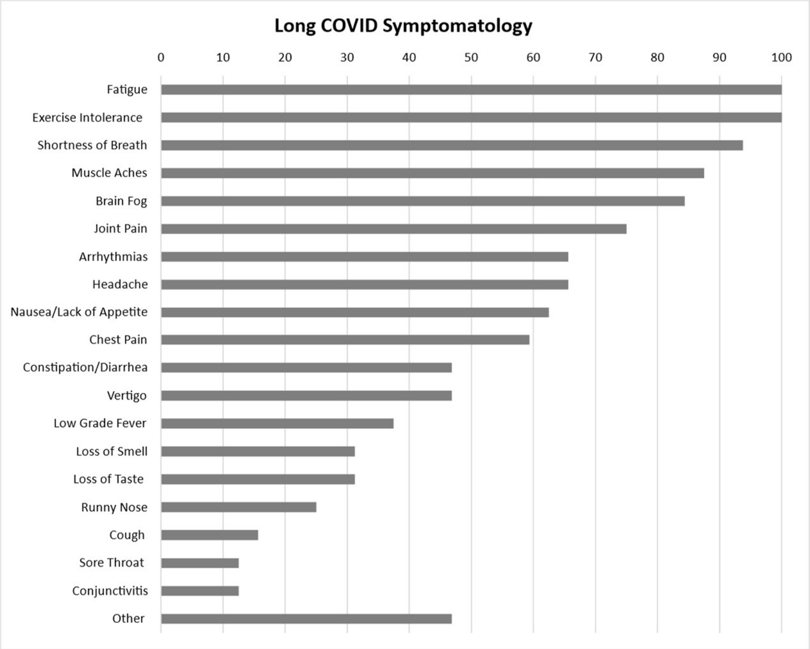


***Figure S1***. *Self-reported symptomatology for participants with Long COVID (n=32) (%).*

| Long COVID (n=32) | n(%) |
|---|---|
| Migraine | 14(44%) |
| Arthritis | 2(6%) |
| Lupus | 0(0%) |
| Mental health condition | 2(6%) |
| Chronic neurological disorder | 0(0%) |
| Peptic ulcer disease | 3(9%) |
| Liver disease | 0(0%) |
| Kidney disease | 1(3%) |
| Heart disease (including congenital) | 2(6%) |
| Family history CVD | 15(47%) |
| Hypertension | 5(16%) |
| Dyslipidemia | 3(9%) |
| Asthma | 6(19%) |
| Chronic Obstructive Pulmonary Disease | 0(0%) |
| Type 1 Diabetes | 0(0%) |
| Type 2 Diabetes | 3(9%) |

***Table S1.*** *Self-reported pre-existing comorbidities for participants with Long COVID (n=32). CVD Cardiovascular Disease.*

| mean±SD | Healthy Controls (n=18) | Long COVID (n=18) | p-value |
|---|---|---|---|
| Peak $\dot{V}O_2$ [ml/kg/min] during exercise | 23.7±5.3 | 18.6±5.3 | <0.001 |
| Extrapolated $\dot{V}O_2$ max [ml/kg/min] | 33.6±7.4 | 29.7±6.6 | 0.02 |
| OUES [ml/min] | 2102.2±530.6 | 1805.2±527.9 | 0.01 |
| $\dot{V}O_2$ at AT [ml/Kg/min] | 16.4±3.3 | 13.1±3.3 | <0.001 |
| VAT [% of predicted] | 56±9 | 48±9 | 0.001 |
| $\dot{V}E/\dot{V}CO_2$ Slope | 25.4±2.7 | 28.3±3.7 | 0.004 |
| $\dot{V}O_2$/WR [ml/min/W] | 9.3±1.1 | 8.4±1.1 | 0.04 |

***Table S2.*** *Cardio-Pulmonary Exercise Test results for 18 participants with Long COVID versus age/sex-matched healthy control participants. $\dot{V}O_2$ (Oxygen Uptake), AT (Anaerobic Threshold), $\dot{V}E/\dot{V}CO_2$ (Ventilation/Carbon Dioxide Production), OUES (Oxygen Uptake Efficiency Slope), $\dot{V}O_2WR$ (Oxygen Uptake Work Rate).*

| n=26 | mean±SD or median[IQR] |
|---|---|
| Number of Intervals | 336±50 |
| Average NN Interval [ms] | 906.8±129.5 |
| TP [ms2] | 1222[2953] |
| VLF [ms2] | 639±485 |
| HF normalised | 50.8±19.8 |
| HF peak [mHz] | 272.1±60.2 |

***Table S3.*** *Additional Heart Rate Variability parameters for participants with Long COVID (n=26). TP (Total Power), VLF (Very Low Frequency), HF (High Frequency).*
